# AI-Powered cellular morphometric biomarkers discovered in needle biopsy of prostatic cancer predict neoadjuvant androgen deprivation therapy response and prognosis: an international multicenter retrospective study

**DOI:** 10.1101/2024.11.17.24317411

**Authors:** Hong Yan, April W. Mao, Dan Li, Manuel Jesús Pérez-Baena, Alejandro Jiménez-Navas, Dawei Wang, Ryan Hong, Weidong Cai, Jesus Pérez-Losada, Kuang-Yu Jen, Sen Wang, Shan Peng, Mary Helen Barcellos-Hoff, Jian-Hua Mao, Yao Fu, Kenneth A. Iczkowski, Shuchi Gulati, Hang Chang

## Abstract

It is imperative to identify patients with prostate cancer (PCa) who will benefit from androgen receptor signaling inhibitors that can impact quality of life upon prolonged use. Using our extensively-validated artificial-intelligence technique: cellular morphometric biomarker via machine learning (CMB-ML), we identified 13 CMBs from whole slide images of needle biopsies from the trial specimens (NCT02430480, n=37) that accurately predicted response to neoadjuvant androgen deprivation therapy (NADT) (AUC: 0.980). Notably, 13-CMB model stratified PCa patients into responder and non-responder groups after NADT treatment in an independent hospital cohort (n=122) that significantly associated with pathologic complete response (p=0.0005), biochemical-recurrence-free survival (p=0.024) and mTOR signaling pathway (p=0.03), suggesting potentially more clinical benefit from mTOR inhibitors in non-responder group. Additionally, genetic and genomic analysis revealed interplay between genetic variants and CMBs on NADT resistance, and provided molecular annotations for CMBs. Overall, prospective clinical implementation of 13-CMB model could assist precision care of PCa patients.

**Significance:** We describe a highly accurate CMB model to predict the therapeutic benefit in prostate cancer patients and uncover the complex interplay between genetic variants and CMBs on NADT resistance. Our model relies only on widely available needle biopsy specimens and provides a robust and cost-effective solution for clinical implementation.

## Introduction

Prostate cancer (PCa) is the most commonly diagnosed non-skin cancer in men in the United States and will be the second-leading cause of cancer-related death in men in 2024 (1), with an estimated 299,010 new cases and 35,250 deaths this year (2). Developing accurate and cost-effective biomarkers to identify men at the greatest risk of poor outcomes following intervention is crucial.

Many patients with localized PCa, i.e., without metastases in other organs or non-regional lymph nodes, can be cured with appropriate management. Depending on the risk stratification using clinical and pathological features (3) and life expectancy, those with low-risk disease may be managed through active surveillance, radical prostatectomy, or radiation therapy (4). In contrast, those with intermediate-high risk may need additional therapies such as androgen deprivation therapy (ADT), androgen receptor signaling inhibitors, and chemotherapy.

Although the effectiveness of neoadjuvant ADT (NADT) was demonstrated in a series of phase-II clinical trials (5–14), both European and American guidelines recommend against the use of NADT prior to radical prostatectomy due to a lack of clinically significant efficacy results and notable side effects (3,15). Other outcomes, such as pathologic complete response rate, varied across cohorts. Correlative studies from window of opportunity trials, however, did show a subgroup of patients that had favorable pathologic responses, and reduced cancer volume at the time of surgery (16–18), with the use of NADT, thus suggesting that patients who responded to, or were resistant to, hormonal therapies could be predicted.

Multidisciplinary cancer research has been essential for precision medicine and personalized therapy of cancer patients, including patients diagnosed with PCa. To allow accurate and cost-effective cancer patient stratification, we have recently developed and validated an artificial intelligence framework for cellular morphometric biomarker (CMB) discovery from whole slide images (WSIs) of tissue histology. In other cancers, CMBs are associated with specific molecular alterations, immune microenvironment, prognosis, or treatment response (19). In this study, we hypothesized that CMBs assessed from PCa can enable precision prognosis and prediction of response to ADT or other therapeutic agents.

## Results

### Retrospective Study Cohorts

The clinical trial cohort (NCT02430480, **Supplementary Table 1**) enrolled thirty-seven patients with intermediate or high-risk PCa, whose targeted biopsies had been obtained before they received six months of NADT plus enzalutamide. A cut point of 0.05 cm3 for residual cancer burden was used to define exceptional responders (ER, n=15) and incomplete and non-responders (INR, n=22) (20,21). Sections of formalin-fixed, paraffin-embedded (FFPE) needle biopsy tissues were stained with hematoxylin and eosin (H&E), and slides were scanned at 20x magnification. The retrospective validation hospital cohort (**Supplementary Table 2**) consisted of 122 needle biopsies (obtained before NADT treatment) from localized primary PCa patients at or above intermediate risk according to National Comprehensive Cancer Network (NCCN) risk classification (22) with complete clinical, pathological, and follow-up information. Sections of H&E stained, FFPE needle biopsy slides were scanned at 20x magnification. The TCGA-PRAD cohort (**Supplementary Table 3**) comprises 396 H&E-stained diagnostic slides from localized primary PCa patients with matching clinical data.

### CMBs accurately predict NADT response in the NCT02430480 trial cohort

CMBs are artificial intelligence-mined imaging biomarkers from WSIs that have demonstrated association with tumor microenvironments, prognosis, and treatment response in different tumor types (23–25) and model systems (26). The predictive CMBs (**Figure 1A** and **1B**) were mined from baseline tissue needle biopsy from patients with intermediate- to high-risk PCa categorized as ER (n=15) or INR (n=22). A total of 256 CMBs were identified, among which the abundance levels of 13 CMBs (i.e., low and high) were significantly associated with treatment response (**Figure 1E** and **1F**, chi-square test, p<0.05; **Supplementary Table 4**). A LASSO regression model based on the abundance levels of these 13 CMBs (13-CMB model) accurately predicted response to NADT (**Figure 1G**, AUC: 0.980; accuracy: 0.892; sensitivity: 0.818; specificity: 1.000), compared to other promising models previously reported (20). Precisely (**Figure 1G** and **1H**), the baseline 4-factor-model (AUC=0.891) consists of four factors: IDC-P (presence of intraductal carcinoma); 10q loss (at least half of chromosome arm 10q deleted hemizygously as determined using the GISTIC algorithm (27)); ERG (immunohistochemical overexpression of nuclear ERG); TP53 (loss-of-function alterations or hotspot mutations to TP53, including copy number loss, as determined by GISTIC). The 4-factor model performance was further improved by including magnetic resonance imaging baseline tumor burden (AUC=0.973) or baseline relative tumor burden (AUC=0.976). Despite these refinements on the 4-factor model, the CMB model continued to perform better (**Figure 1G** and **1H**) in comparison, thus making the CMB model an accurate option for patient selection.

**Figure 1.**
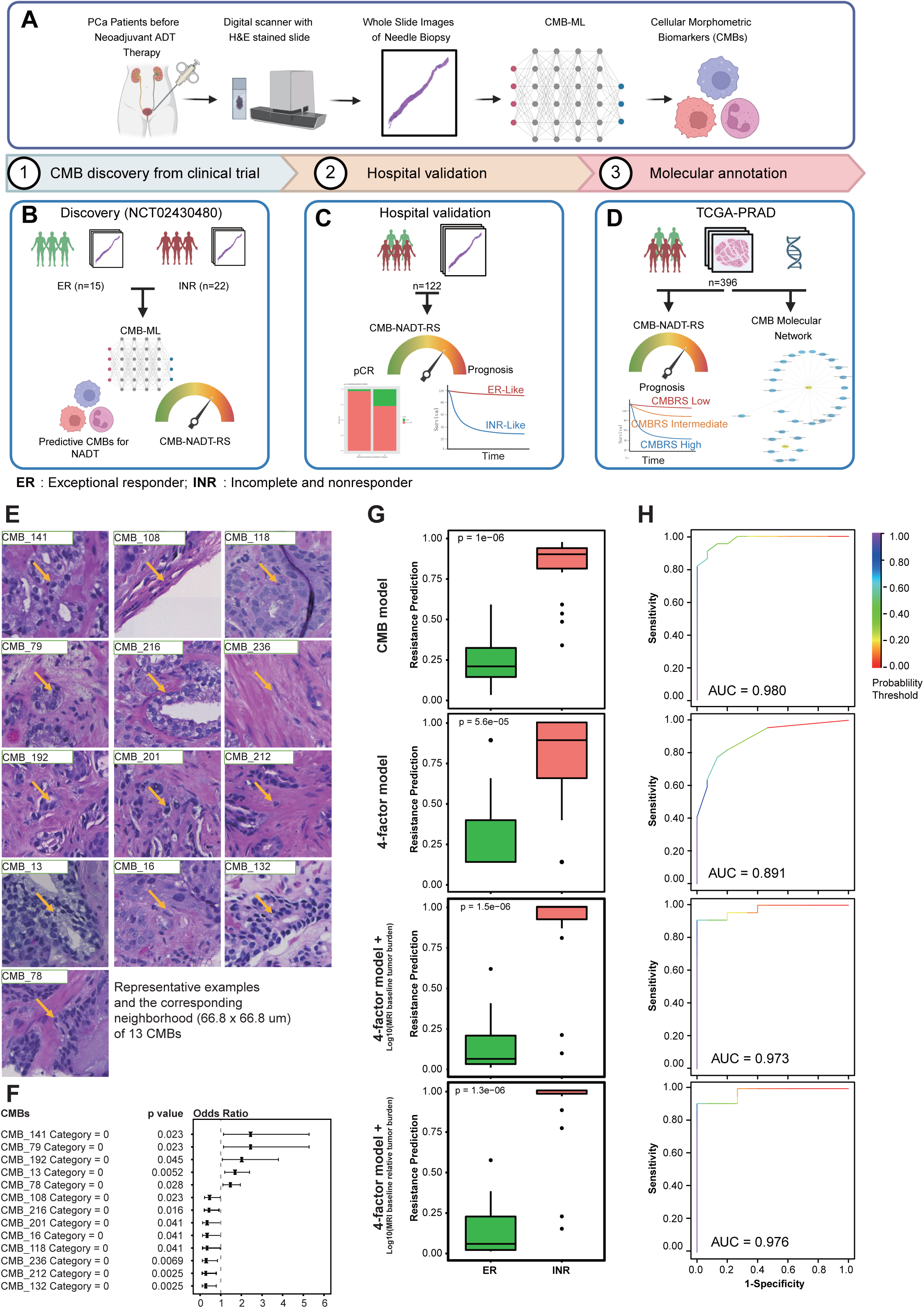
(**A-D**) Study design. (**A**) CMB-ML pipeline for the discovery of CMBs from needle biopsy of NADT patients; (**B**) Discovery cohort; (**C**) Hospital validation cohort; (**D**) Molecular annotation. (**E**) Representative examples of CMBs. (**F**) Association of individual CMB with treatment resistance; (**G**) Predictive probability between ER and INR groups across different models, where the 4-factor model is based on IDC (presence of intraductal carcinoma); 10q loss (at least half of chromosome arm 10q deleted hemizygously as determined using the GISTIC algorithm); ERG (overexpression of nuclear ERG determined by immunohistochemistry); TP53 (loss-of-function alterations or hotspot mutations to TP53, including copy number loss, as determined by GISTIC); (**H**) Receiver operating characteristic (ROC) curve across different models. The p values in (**F**) were obtained using logistic regression, and the p values in (**G**) were obtained using Non-parametric Mann-Whitney tests. ***Abbreviations***: ER: Exceptional responder; INR: Incomplete and non-responder. Panels (A-D) are created with BioRender.com.

### CMBs stratified PCa patients treated with NADT from an independent hospital cohort into groups with distinct treatment responses and prognosis

To test the predictive value of the 13-CMB model, an independent cohort of 122 localized PCa patients treated with NADT was assessed (**Figure 2A**). 13-CMB model classified patients into ER-like (n=22) and INR-like (n=100) groups. The ER-like group had significantly better rates of pathologic complete response (p=0.0005, **Figure 2B**), and significantly better biochemical-recurrence-free survival (p=0.024, **Figure 2C**). We next applied the 13-CMB model on the TCGA-PRAD cohort of 396 localized primary PCa patients with diagnostic slides and classified them into ER-like (n=252) and INR-like (n=144) groups (**Figure 2 D-F**). In this cohort, the ER-like group had significantly better progression-free survival (PFS) (**Figure 2E**, p=0.0017) than patients in the INR-like group. Lastly, we constructed a cellular morphometric biomarker risk score (CMBRS) based on 13 CMBs that stratified the TCGA-PRAD cohort into CMBRS-low, intermediate, and high groups with significant prognostic value after adjusting for important clinical factors such as stage, Gleason score, PSA at the time of diagnosis, and age (**Figure 2 G-I**). Our findings confirm that the 13-CMB model carries both predictive and prognostic value in patients with localized PCa treated with/without NADT.

**Figure 2.**
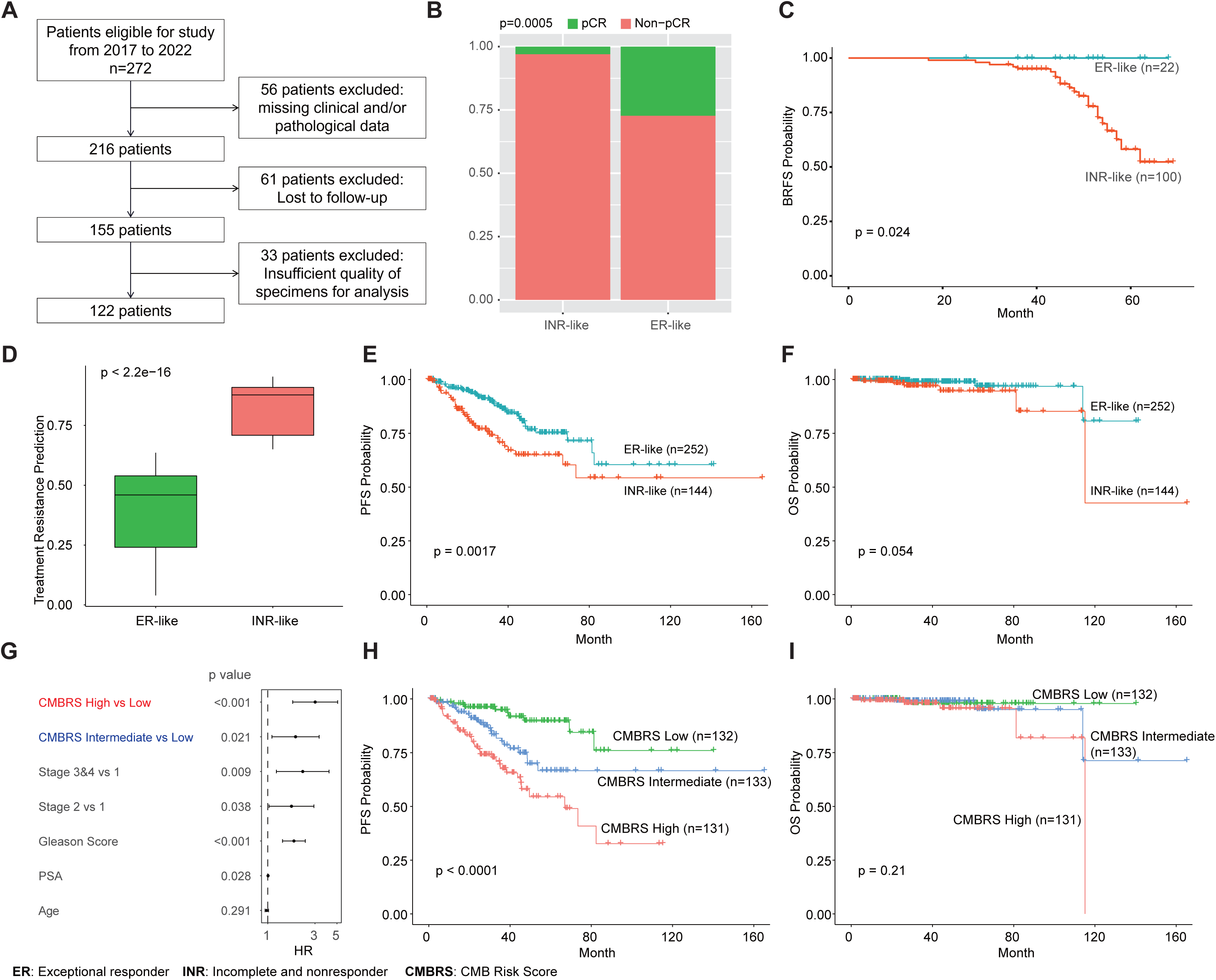
(**A**) Patient inclusion chart for the hospital validation cohort. (**B**) 13-CMB model is significantly associated with NADT response. (**C**) 13-CMB model is significantly associated with biochemical recurrence-free survival. (**D-F**) 13-CMB model stratifies TCGA-PRAD patients into groups with significantly different resistance risk score (**D**), and progression-free survival (**E**), but no difference in overall survival (**F**). (**G-I**) Re-optimization of 13 CMBs in the TCGA-PRAD cohort provides independent and significant prognostic value after adjusting for clinical factors (**G**) that stratify TCGA-PRAD patients into groups with significantly different progression-free survival (**H**) despite no difference in overall survival (**I**).

### The interplay between genetic variants and cellular morphometric biomarkers contributes to NADT resistance

Germline genetic single nucleotide polymorphisms (SNPs) have been identified by genome-wide association studies and found to be associated with PCa (28) and the treatment response to ADT (29). However, the interplay between genetic variants and cellular morphometric architecture on treatment response remains unknown. To explore the potential relationships between genetic variants and CMBs as well as their synergistic effect on the response to NADT, we studied the 146 risk SNPs profiled in the NCT02430480 cohort. Among the 13 CMBs, 12 were significantly associated (Mann-Whitney non-parametric, p<0.05) with 34 SNPs (**Figure 3A**). Interestingly, through mediation analysis, we identified that the interplay between 4 SNPs and 2 CMBs (rs61890184 and CMB 236; rs56232506 and CMB 216; rs130067 and CMB 216; rs10009409 and CMB 216) significantly contribute to the individual response to NADT (total effect p < 0.05; average causal mediation effects (ACME) p < 0.05; **Figure 3B**). Unsurprisingly, the four genes associated with above four SNPs (rs10009409: *COX18*; rs130067: *CCHCR1*; rs56232506: *TNS3*; rs61890184: *PPFIBP2*) provide significant and independent prognostic value in TCGA-PRAD cohort, after adjusting for CMBs and key clinical factors such as stage, Gleason score, PSA level at diagnosis and age (**Figure 4 A-C**). The multimodal signature (combining the SNP-associated four genes and 13 CMBs) improved stratification in the TCGA-PRAD cohort, especially for identifying the patient subgroup with the poorest PFS and hence worse prognosis (**Figure 4 D-F**). Our findings uncover a complex interaction between genetic variants and cellular morphometric architecture that together help predict response to NADT.

**Figure 3.**
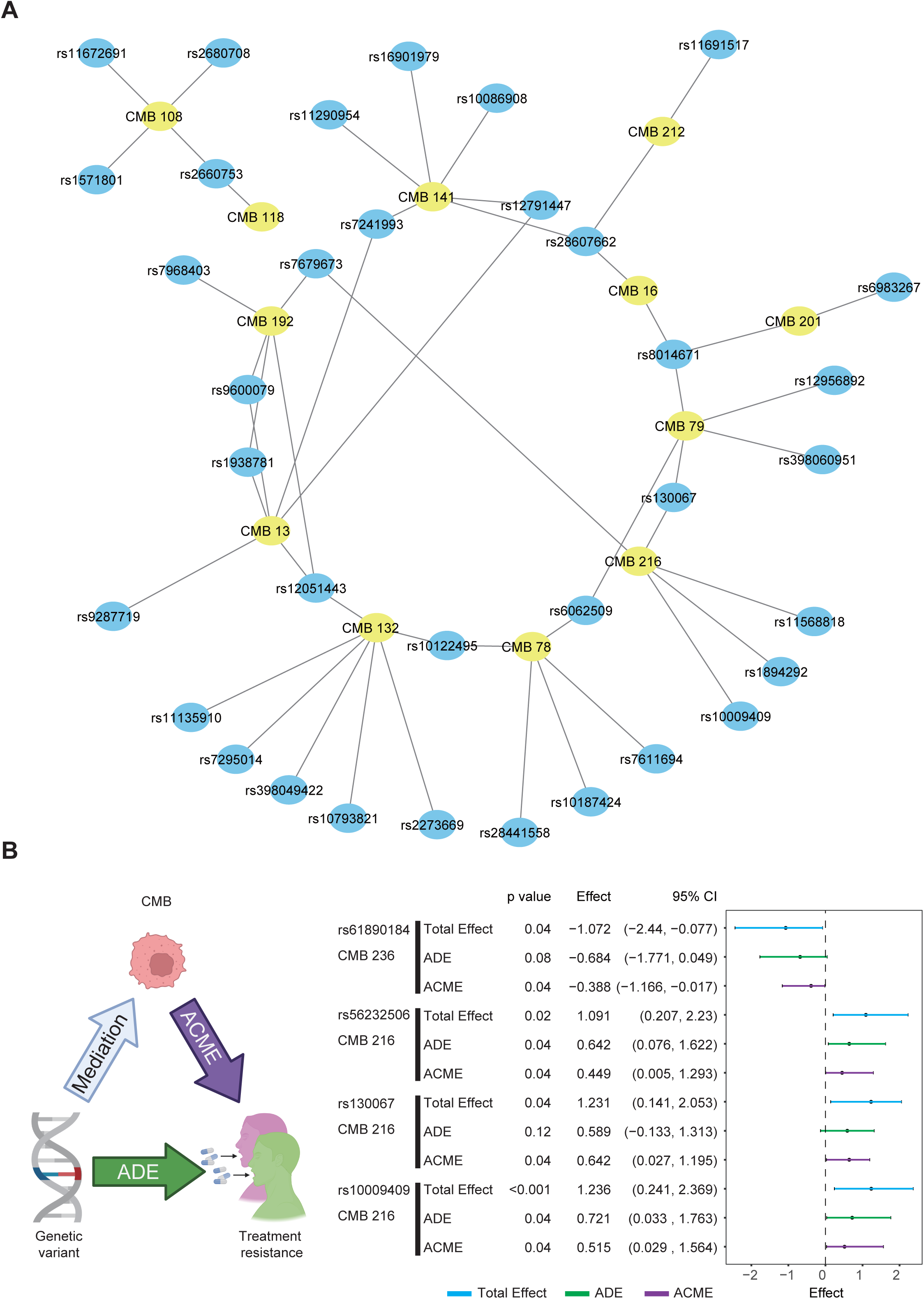
(**A**) Association network between CMBs and risk SNPs previously identified by GWAS projects as having significant associations with prostate cancer; (**B**) Four genetic variants and two CMBs that mediate NADT treatment resistance together (ADE: Average Direct Effects; ACME: Average Causal Mediation Effects). Figure 4B is created with BioRender.com.

**Figure 4.**
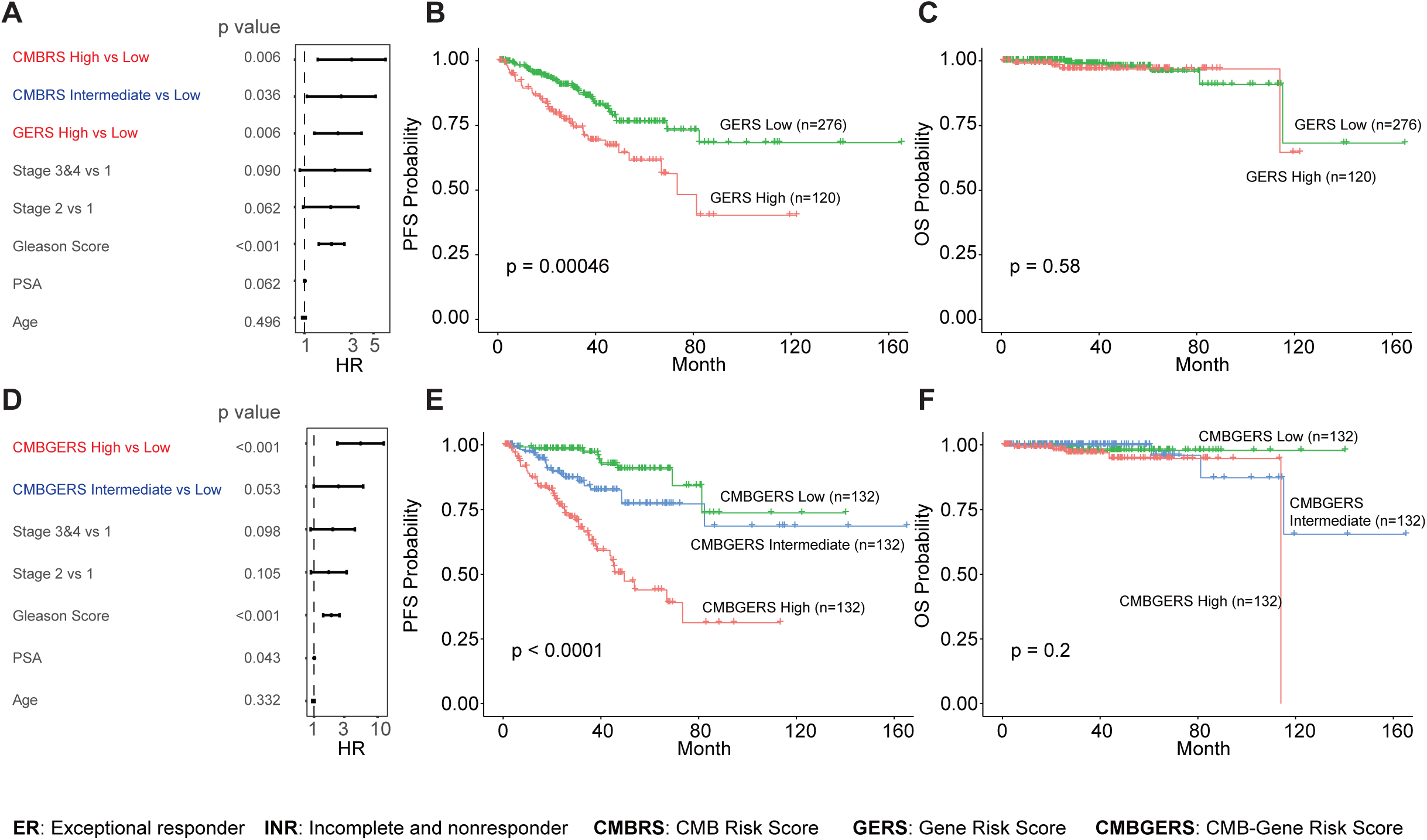
(**A-C**) Four genes associated with four SNPs (identified by mediation analysis) that provide independent and significant prognostic value, after adjusting for clinical factors and CMBRS (**A**). These stratify TCGA-PRAD patients into groups with significantly different progression-free survival after adjusting for clinical factors (**B**), despite no difference in overall survival (**C**). The combination of 13-CMB and 4-Gene signatures provides independent and significant prognostic value (**D**) that stratifies TCGA-PRAD patients into groups with significantly different progression-free survival (**E**) despite no difference in overall survival (**F**).

### CMBs are significantly associated with tumor microenvironment

PCa has a distinct tumor microenvironment (TME) consisting of stromal cells, immune cells, and a dense extracellular matrix. The TME has been shown to play a role in determining survival, therapeutic response, and metastasis (30). The TCGA-PRAD cohort enables efficient assessment of the association between CMBs and the previously inferred (i.e., using RNA-seq) TME constituents (**Supplementary Figure 2A**). Interestingly, CMB 141, an unfavorable CMB in NADT treatment response (**Figure 1F**, Odds Ratio > 1), was seen to be significantly and positively associated with immune cell infiltration in the TME, including CD4+ T cells, regulatory T cells, mast cells, and M2 macrophages. At the same time, CMB 236, a favorable CMB for NADT treatment response (**Figure 1F**, Odds Ratio < 1), was seen to be significantly and negatively associated with immune cell infiltration in the TME.

### CMBs are significantly associated with genome instability

As a heterogeneous multifocal cancer, localized PCa has signs of genomic instability (31,32) that are associated with recurrence and progression to aggressive cancer (33). Using the resources provided in the TCGA-PRAD cohort, we revealed a significant association between CMBs and genomic instability (GI) (**Supplementary Figure 2B**). Specifically, CMB 141, an unfavorable CMB in NADT treatment response (**Figure 1F**, Odds Ratio > 1), was significantly associated with elevated genomic instability in terms of aneuploidy score, microsatellite instability (MSI) MANTIS score, and fraction of genome altered. CMB 212 and 236, favorable CMBs for NADT treatment response (**Figure 1F**, Odds Ratio < 1), were significantly associated with lower genomic instability regarding tumor mutation burden, nonsynonymous mutation count, and MSIsensor score.

### CMBs are significantly associated with essential molecular functions

To gain insight into molecular annotation underlying CMBs, we identified genes significantly associated with individual CMB in the TCGA-PRAD cohort, performed Gene Ontology (GO) functional enrichment analysis on biological process (BP), cellular components (CC), molecular function (MF), and Kyoto Encyclopedia of Genes and Genomes (KEGG) pathway, and explored the molecular annotation underlying CMBs through the CMB-BP-Network (**Figure 5A**), CMB-CC-Network (**Supplementary Figure 3**), CMB-MF-Network (**Supplementary Figure 4**) and CMB-KEGG-Network (**Supplementary Figure 5**). Interestingly, many of the CMBs (e.g., CMB 16, CMB 108, CMB 118, CMB 201, CMB 212, CMB 236) co-registered with cell cycle-related biological processes, which play an essential role in drug resistance such that inhibition of prostate cell proliferation helps overcome the resistance to AR inhibitors (34). Other CMBs are associated with distinct functional groups (e.g., CMB 132, CMB 216), revealing potentially different molecular functions underlying these CMBs.

**Figure 5.**
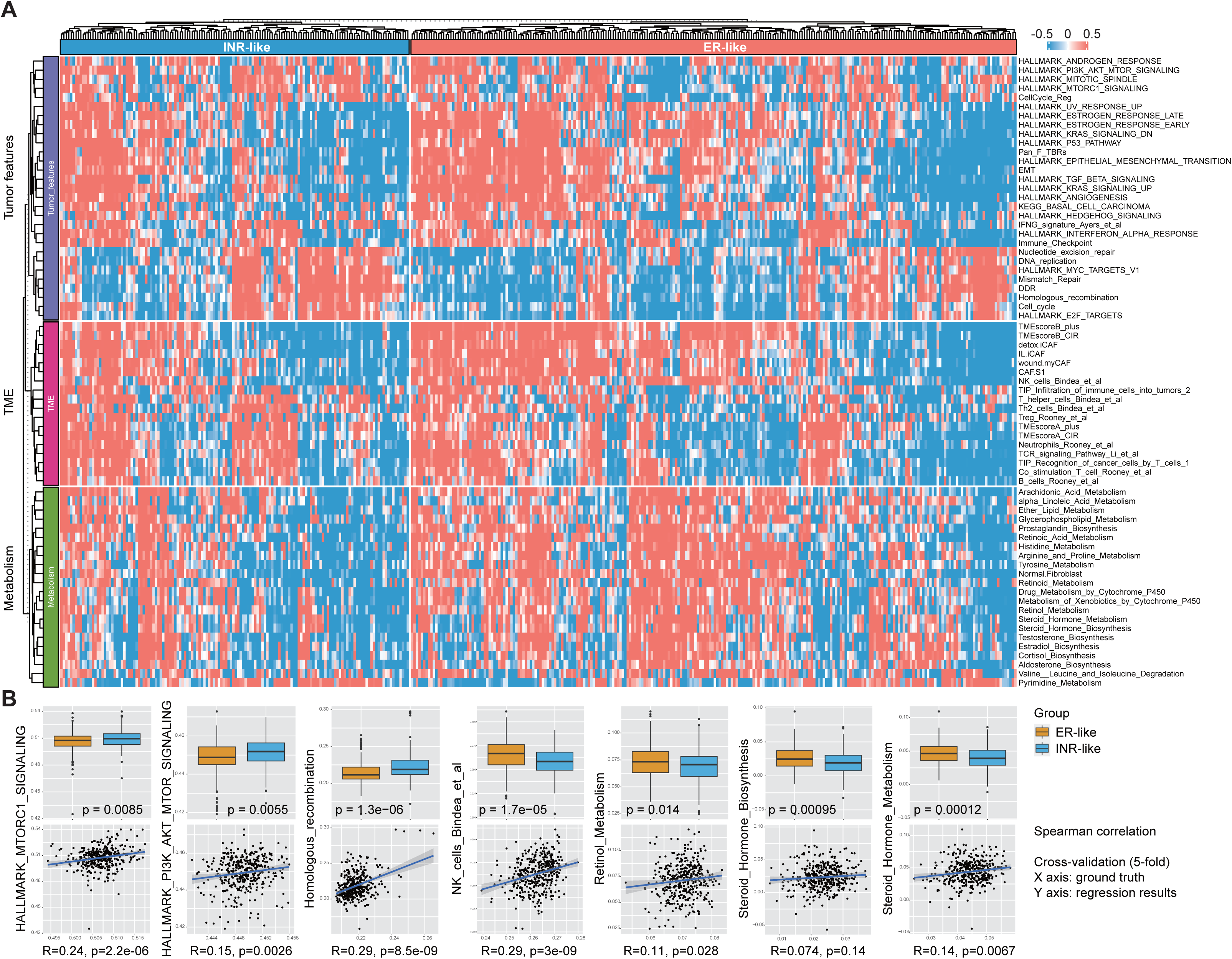
Association between ER- or INR-like groups and signatures related to tumor features, tumor microenvironment (TME), and metabolism (**A**); Examples of significantly different signature scores between ER- or INR-like groups (**B**, top pane); and prediction of these signature scores on hold-out samples during 5-fold cross-validation (**B**, bottom pane).

### ER-like and INR-like patients have distinct signatures related to tumor features, TME, and metabolism

In the TCGA-PRAD cohort, the distinction of molecular features between ER-like and INR-like groups was characterized by tumor functional state feature-related signatures; TME-related signatures, including immune and stromal components; and metabolic reprogramming signatures (**Figure 5A and 5B, Supplementary Figure 7**). Specifically, 29 signatures (including PI3K_AKT_MTOR_SIGNALING, cell cycle, Homologous recombination repair (HRR), DNA damage repair (DDR), Endothelial-to-Mesenchymal transition (EMT), Mismatch repair (MMR), and MTORC1_SIGNALING) were used to describe the functional states of tumor cells. HRR signature (**Figure 5B** top pane, p=1.3e-06), PI3K_AKT_MTOR (**Figure 5B** top pane, p=0.0055), and DDR signature (**Supplementary Figure 7**, p=1.3e-05) had higher GSVA scores in the INR-like group compared to the ER-like group. In comparison, the signature of the ESTROGEN_RESPONSE functional states of tumor cells (**Supplementary Figure 7**, p=0.0041) had higher GSVA scores in the ER-like group. We observed marked upregulation of anti-tumor immune components, such as natural killer cells and T regulatory cells (**Figure 5B** top pane, p=1.7e-05), suggesting immune escape components were distinctly regulated in the INR-like group. Metabolic reprogramming also differed significantly between the ER-like and INR-like groups. We analyzed the metabolic pathways obtained from the KEGG database and observed that steroid hormone metabolism (**Figure 5B** top pane, p=0.00012), and steroid hormone biosynthesis (**Figure 5B** top pane, p=0.00095) downregulated in the INR-like group. Unsurprisingly, the 13 CMBs were predictive of many of these signature scores, including HRR (**Figure 5B** bottom pane, R=0.29, p=8.5e-09), natural killer cells (**Figure 5B** bottom pane, R=0.29, p=3e-09) and DDR (**Supplementary Figure 7**, R = 0.27, p=8.6e-06). In addition, using these pre-built CMB-signature models from the TCGA-PRAD cohort, we can predict the signature scores in the independent hospital cohort, which show significant differences between the ER-like and INR-like patient groups and are consistent with our observations in the TCGA-PRAD cohort (**Supplementary Figure 8, Supplementary Table 5**).

### INR-like patients have more robust activation of the mTOR signaling pathway and are more sensitive to mTOR inhibitor

Applying the pre-built CMB-signature models from the TCGA-PRAD cohort on a subset of patients in the independent cohort (n=20) with matching specimens before and after NADT treatment revealed the impact of NADT treatment on various molecular signatures (**Supplementary Table 6)**. Specifically, in both ER-like (n=10) and INR-like (n=10) groups, NADT treatment led to increased NK cells (**Figure 6B**; ER-like: p=4.1e-05; INR-like: p=2.8e-6), increased EMT (**Figure 6C**; ER-like: p=4.1e-05; INR-like: p=2.8e-6), and defected homologous recombination repair (**Figure 6D**; ER-like: p=0.00016; INR-like: p=5.7e-6). Interestingly, the mTOR activity seems not to be affected by NADT treatment (**Figure 6A**; ER-like: p=0.67; INR-like: p=0.065), and it remains higher in the INR-like group regardless of NADT treatment. This observation was consistent with the higher protein expression related to mTOR pathway in the TCGA-PRAD INR-like group (**Figure 6E**; p-mTOR: p=0.15; p70S6K: p=1.6e-05) and suggested that the INR-like group is significantly more sensitive to mTOR inhibitors (**Figure 6G**-**8H**; Rapamycin: p=8.3e-05; Temsirolimus: p=0.57). Notably, the substantially higher mTOR activity in the INR-like group was validated using immunohistochemical (IHC) staining in the independent hospital cohort (**Figure 6I**-**8K**, p-mTOR, p=0.063; **Figure 6M**-**8O**, p70S6K, p=0.03; **Supplementary Table 7**). The strong and significant correlation between the predicted mTOR signaling score and the protein expression of p-mTOR (**Figure 6L**, R=0.66, p=0.0015) and p70S6K (**Figure 6O**, R=0.68, p=0.0013) confirmed the predictive power of the prebuilt CMB-signature models.

**Figure 6.**
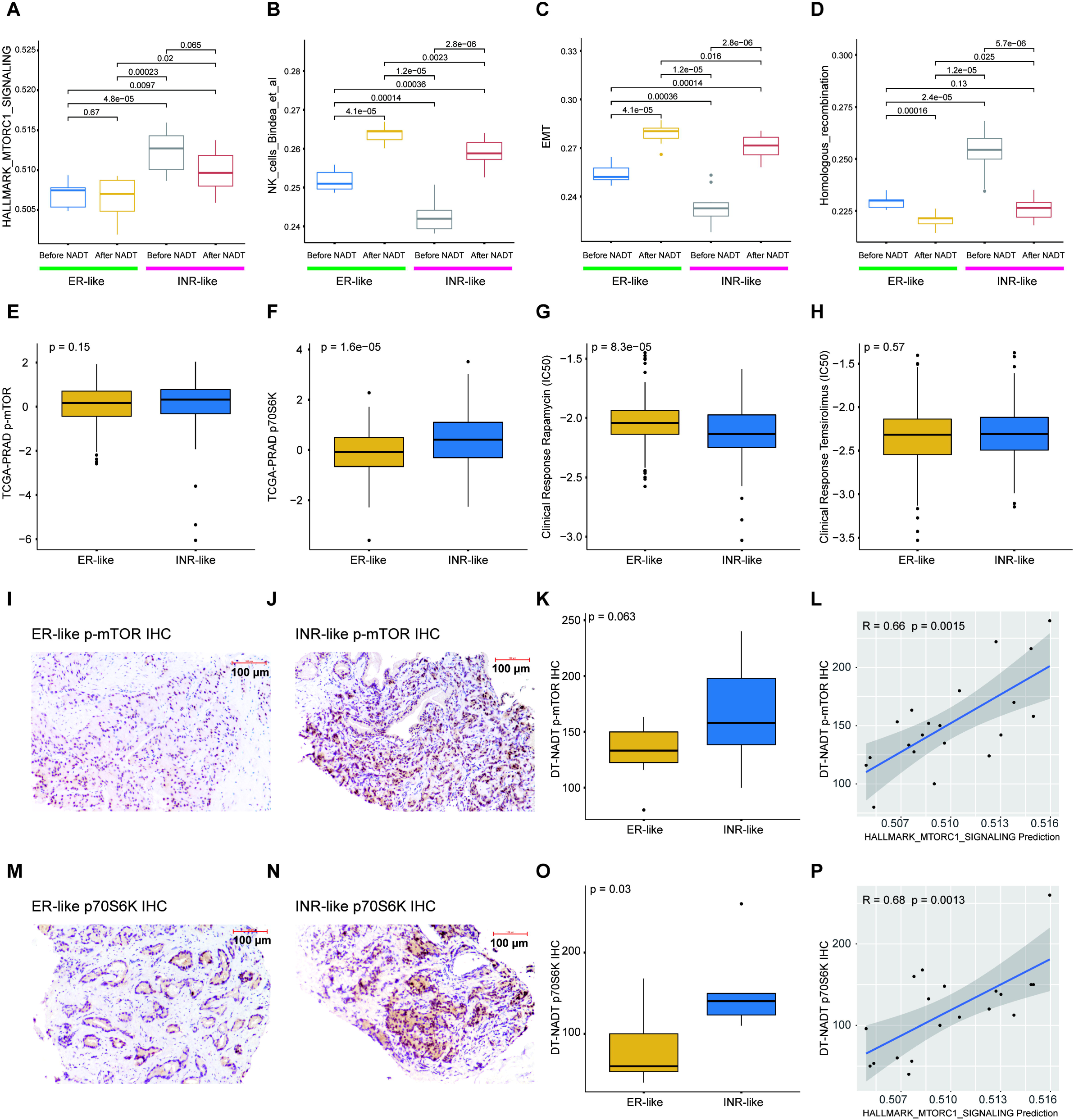
Predicted (**A**) HALLMARK_MTORC1_SIGNALING, (**B**) NK_cells, (**C**) EMT, and (**D**) Homologous_recombination scores before and after NADT treatment in the independent hospital cohort, using CMB-Signature models pre-built from TCGA-PRAD cohort. (**E**-**F**) Protein expression of p-mTOR and p70S6K in TCGA-PRAD ER-like and INR-like tumors. (**G**-**H**) Sensitivity prediction of mTOR inhibitors (i.e., rapamycin and temsirolimus) between TCGA-PRAD ER-like and INR-like tumors based on RNA-seq data using pRRophetic in TCGA-PRAD cohort, where rapamycin shows significantly higher sensitivity in the INR-like tumors as indicated by lower predicted IC50 values. (**I**-**K**) Immunohistochemical (IHC) staining shows the tendency of more p-mTOR in INR-like tumors in the independent hospital cohort, and (**L**) the p-mTOR protein expression is strongly and significantly correlated with the predicted HALLMARK_MTORC1_SIGNALING score. (**M**-**O**) Immunohistochemical (IHC) staining shows significantly more p70S6K in INR-like tumors in the independent hospital cohort, and (**P**) the p70S6K protein expression is strongly and significantly correlated with the predicted HALLMARK_MTORC1_SIGNALING score. Scale bar = 100 µm, p values in (**A**-**H**, **P**, **O**) were obtained using non-parametric Mann-Whitney tests, and p values in (**L**, **P**) were obtained using the Spearman correlation.

## Discussion

Intense NADT trials have yielded lower recurrence rates among PCa patients with minimal residual disease after treatment (5–14). Genomic and histologic features associated with NADT treatment resistance at baseline have recently been identified (20); here, we developed and validated robust artificial intelligence-powered CMB predictive biomarkers of NADT response. Discovery using whole slide images of needle biopsies from a clinical trial cohort identified 13 CMBs employed in a predictive model, the 13-CMB model. This model was validated in two independent cohorts to accurately predict both response to NADT and prognosis. We also describe the interplay of the CMBs with genetic variants towards NADT resistance, and association with distinct molecular alterations and the TME. Furthermore, we constructed predictive models using 13 CMBs towards various molecular signatures, validated the predictive power and our findings on the mTOR signaling pathway through IHC staining, and revealed potential benefits from mTOR inhibition in patients resistant to NADT. In addition, the predicted changes of specific molecular signatures, including NK cells, EMT and Homologous recombination before and after NADT treatment are consistent with our current understanding of the impact of ADT on DNA repair (35), EMT (36), and immune system response (37). The application of CMBs may provide an accurate method for personalized management of PCa, whereas the association of CMB with specific molecular features may help with developing an understanding of its biology, thus facilitating future drug development.

Current guidelines (3) include clinical and pathologic features, such as PSA level, to define risk classes in prostate cancer. Prior studies have shown clinical factors such as baseline PSA and PSA dynamics and extent of bone involvement as predictors of response to ADT (38). Molecular features such as loss of chromosome 10q (containing *PTEN*) and *TP53* alterations, as well as nuclear ERG expression and the presence of intraductal carcinoma of the prostate have been shown to be predictive of poor outcomes (20). Androgen receptor mutations are frequently seen in patients with metastatic castrate-resistant PCa (39) and prior studies have shown these mutations may even contribute to resistance to androgen receptor signaling inhibitors such as enzalutamide and abiraterone (40). Additional mutations such as *ATM, BRCA,* and *TP53* may confer resistance to these drugs and thus lead to poor outcomes as well (40). Baseline tumor volume estimation using a multi-parametric MRI was predictive of response to NADT as well. The CMB model we describe here was shown to perform better than the described models that incorporated an MRI, presence of intraductal carcinoma, 10q loss, nuclear ERG overexpression, or altered *TP53*.

Previously, genome-wide association studies have identified >100 common SNPs that may be associated with susceptibility to PCa (41). Genetic polymorphisms associated with the efficacy of ADT have also been described (29). In this study, we reveal the potential interplay between genetic variants and cellular morphometric architecture on NADT treatment response and describe a multimodal signature that combines the four SNP-associated genes and 13 CMBs with an increased power to predict prognosis. This is the first effort where the causal effects between genetic polymorphic changes and cell morphometric architecture on NADT response are described.

Genomic instability (31,32) is associated with localized PCa recurrence and progression to aggressive cancer (33). It is reported that a high tumor mutational burden (TMB) and MSI-high status are predictive of response to immune checkpoint blockade in patients with PCa (42,43). In our analysis, we discover that the majority of CMBs associated with favorable response to NADT treatment are associated with lower TMB and/or MSI. In contrast, the majority of CMBs associated with unfavorable NADT response are related to elevated TMB and/or MSI, suggesting that this group of patients may have a better response if treated additionally with immune checkpoint inhibitors. Thus, certain CMBs could be used in conjunction as biomarkers that predict response to immunotherapy with or without ADT (44–46).

The TME includes immune and non-immune components that have been shown to contribute to disease prognosis (47). In our analyses, we report the association of TME immune elements (CD4+ T cells, regulatory T cells, and mast cells) with a CMB that associates with poor response to ADT. Previous studies have shown mast cells to be pro-tumorigenic (48). There have been conflicting data on the relationship between infiltration of lymphocytes and survival in prostate cancer. However, studies suggest CD4+ regulatory T cells are associated with a worse prognosis (49).

In addition to predicting response to ADT, we also show that certain CMBs may be able to predict molecular signatures and the response to drugs targeting specific pathways, including the mTOR pathway and DDR pathway. The abnormal regulation of mTOR has been extensively reported within human carcinomas from several different origins (50), and the combination of ADT and mTOR inhibition may be of therapeutic value in PCa treatment (51). We discovered (proteomics in TCGA-PRAD) and validated (IHC in independent hospital cohort) higher mTOR activity in the INR-like group compared to the ER-like group, which is consistent with a previous report of mTOR signaling as a resistance mechanism for PCa ADT therapy (52). Moreover, we revealed higher sensitivity (estimated by IC50 score) to mTOR inhibitor (rapamycin) of the INR-like group compared to the ER-like group, suggesting potential benefit from mTOR targeted therapy for patients in the INR-like group. Although multiple studies have shown no clinical benefit from metformin in prostate cancer (53,54), there was a subgroup of patients with high-volume disease in the latter trial, where clinical benefit was described. These findings align with what we describe here in that we see potential benefits from mTOR pathway inhibition in the INR-like group, warranting further evaluation of drugs to block this pathway in this specific subgroup of patients.

DNA–damage repair (DDR) pathway alterations are detected in about 20% of patients with prostate cancer and are associated with improved sensitivity to poly(ADP ribose) polymerases (PARP) inhibitors. Several mechanisms can be activated to repair damaged DNA, one of the most crucial ones being homologous recombination repair (HRR), with the breast cancer genes (BRCA1 and BRCA2) playing a pivotal role. Recently, several clinical trials have reported on the benefit of PARP inhibitors in these patients (55–57). We discovered that DDR signature has higher activity (estimated by GSVA score) in the INR-like group compared to the ER-like group, a potential benefit from DDR-targeted therapy for patients in the INR-like group (58–62) and also warrants evaluation in a prospective setting.

Steroid hormones, particularly androgens, play an essential role in both the development of benign prostatic hyperplasia and the stimulation of PCa growth (63), and genes associated with steroid hormones predict the PCa prognosis (64). Upregulation of cholesterol and steroid hormone biosynthesis in PCa cells, driving them into androgen receptor targeted therapy resistance. Blocking both pathways may, therefore, be a promising approach to overcome resistance to androgen receptor deprivation therapies in PCa (65), which may explain the ADT sensitivity of patients in the ER-like group, given its elevated activity in steroid hormone biosynthesis.

This study has several limitations. First, the clinical trial cohort for 13-CMB model development was relatively small. Second, the hospital validation was performed retrospectively with only one center involved. Third, the treatment information in the TCGA-PRAD cohort is incomplete. Nevertheless, our findings on both the 13-CMB model and its underlying molecular association warrant future evaluation in larger multicenter cohorts and in a prospective clinical trial. However, our findings demonstrate that the predictive power of the CMB model remains robust even when validated in a heterogeneous dataset such as TCGA-PRAD, which includes variability in clinical practices and incomplete standardization of treatment history. This highlights the model’s ability to generalize across diverse clinical settings, suggesting strong potential for real-world application. The ability to achieve significant results under these conditions supports the resilience and practicality of our AI-driven approach, underscoring its suitability for implementation in diverse and less controlled clinical environments.

In conclusion, we developed the 13-CMB model based on a needle biopsy H&E that may accurately predict NADT treatment response and prognosis in localized PCa. This model shows associations with TME composition, markers of genome instability, and important biological pathways that suggest the underlying molecular mechanisms for its predictive power. We also show the predictive power of the 13-CMB model, including validation of prediction of mTOR activity, where we confirmed the elevated mTOR activity in the INR-like group and revealed the increased sensitivity and potential therapeutic benefit from mTOR inhibitors in the INR-like group. Our study highlights a novel advancement of AI in digital pathology through the unification of robust predictive power with biological interpretability, molecular mechanisms, and treatment optimization potentials, which the classic deep learning systems (66) can rarely offer. Thus, it warrants future prospective validation in larger cohorts.

## Methods

### Retrospective Study Design

The study design is illustrated in **Figure 1 (A-D)**. Specifically, the CMB-ML pipeline was applied on WSI of needle biopsy (**Figure 1A**) from 37 patients enrolled in a clinical trial (NCT02430480) for CMB identification and model construction (**Figure 1B**). The independent validation hospital cohort of 122 localized primary PCa patients at or above intermediate risk according to NCCN risk classification (22) with needle biopsy, complete clinical, pathological, and follow-up information between 2017 and 2022 was retrospectively retrieved from the Nanjing Drum Tower Hospital (**Figure 1C**). The entire cohort of 272 patients included 150 cases excluded due to missing clinical and/or pathological data (56 cases), loss to follow-up (61 cases), and insufficient quality of specimens for analysis (33 cases). Patients were followed up through January 2024. The validation study was independently carried out at the Nanjing Drum Tower Hospital, and the study was approved by the Ethics Board of the Nanjing Drum Tower Hospital with a waiver of informed consent. Clinical, epidemiological, and histopathologic features are summarized in **Table 1**. The TCGA-PRAD cohort (**Figure 1D**), consisting of 396 H&E-stained diagnostic slides from localized primary PCa patients with matching clinical data, was used to evaluate the prognosis and molecular association of the CMB model.

### CMB identification and predictive model construction

Based on the stacked predictive sparse decomposition (67) technique and our cellular morphometric biomarker by machine learning (CMB-ML) pipeline (68–70), we defined 256 CMBs from cellular objects extracted from the WSI of H&E stained tissue histology sections from the needle biopsies of 37 patients enrolled in a clinical trial (NCT02430480). In the CMB-ML pipeline, we used a single network layer with 256 dictionary elements (i.e., CMBs) and a sparsity constraint of 30 at a fixed random sampling rate of 1000 cellular objects per WSIs from the cohort. The pre-trained SPSD model reconstructed each cellular region as a sparse combination of pre-defined 256 CMBs and thereafter represents each patient as an aggregation of all delineated cellular objects belonging to the same patient.

The predictive effect of high or low levels of each CMB on NADT treatment response (i.e., INR and ER) was assessed by chi-square test, where the NCT02430480 cohort was divided into two groups (i.e., CMB-high and CMB-low groups) based on each CMB with cut-off point optimized towards minimized p-value during the chi-square test. The set of CMBs with p-value < 0.05 was selected as a predictive signature for LASSO (Least Absolute Shrinkage and Selection Operator, glmnet package in R, Version 4.1-4) regression model construction towards NADT treatment response (i.e., INR and ER). The model parameter (i.e., lambda and coefficients) was optimized using cross-validation (**Supplementary Figure 1A and B**), and the cut point at 64.2% on estimated probability was optimized by bootstrapping strategy (80% sampling rate with 100 iterations) on the Youden index (cutpointr package in R, Version 1.1.2).

### Exploration of the underlying association between genetic variants and CMBs and their interplay in NADT treatment resistance

Mann-Whitney non-parametric test was used to evaluate a significant association between CMBs and 146 risk SNPs related to PCa provided by the NCT02430480 cohort (p<0.05). CMB-SNP-Network was then constructed and visualized in Cytoscape (version 3.8.2). Mediation analysis, a statistical model to determine whether the relationship between two variables (e.g., genetic variant and NADT treatment resistance) is mediated through a third variable (e.g., CMB), was performed using a mediation package (version 4.4.7) and visualized using the ggplot2 package (version 3.2.1) in R (version 3.6.0).

### Exploration of the underlying association between tumor microenvironments (TMEs) and CMBs in TCGA-PRAD

The TME (i.e., abundances of member cell types in a mixed cell population) was assessed using ConsensusTME (version: 0.0.1.9000) (71). The association between CMBs and TMEs was calculated by Spearman rank correlation, and represented by a heatmap (ComplexHeatmap package in R, version 3.18).

### Exploration of the underlying association between genomic instability and CMBs in TCGA-PRAD

Genomic instability in terms of aneuploidy score and fraction of genome altered, mutation counts, mutation burden, and prognosis (i.e., overall survival and progression-free survival) were downloaded from cBioPortal, and the association between CMBs and genomic instability parameters was calculated by Spearman correlation, and represented by a heatmap (ComplexHeatmap package in R, version 3.18).

### Exploration of underlying molecular associations of CMB in TCGA-PRAD

The CMB-Enrichment-Network study was performed based on the following steps: (1) significantly CMB-associated genes were selected per cancer type per CMB (Spearman correlation, |correlation coefficient|>0.20 and p<0.05, R version 3.6.0); (2) Enrichment analysis (i.e., BP/Biological Process, CC/Cellular Component, MF/Molecular Function, and KEGG/ Kyoto Encyclopedia of Genes and Genomes) was performed (clusterProfiler package in R, version 4.1.0) on CMB-associated genes per CMB; and (3) CMB-BP-Network, CMB-CC-Network, CMB-MF-Network and CMB-KEGG network were then constructed and visualized in Cytoscape (version 3.8.2).

### Evaluation of the TME-related signature scores in TCGA-PRAD patients

After reviewing previously published studies (72–77), the Molecular Signatures Database (MSigDB; http://www.gsea-msigdb.org/gsea/msigdb/index.jsp), and the Reactome pathway portal (https://reactome.org/PathwayBrowser/), we identified relevant biomarker genes for tumor, immune, stromal, and metabolic reprogramming signatures. The 61 TME-related signature as well as the source of each signature were included in this study. The signature score of each TME-related signature was calculated per sample using gene set variation analysis (GSVA) (‘GSVA’ package, version 1.46.0). Except for particular indications, heatmap visualization was achieved using the R package ggplot2 (version 3.4.1).

### Evaluation of other biological pathway enrichment scores in TCGA-PRAD patients

Human metabolism-related pathways were obtained from the KEGG database (https://www.genome.jp/kegg/). A previously published study retrieved 86 human metabolism-related pathways and ten oncogenic signatures containing an HR signature. GSVA was performed to calculate the enrichment score of each signature for each sample. To identify the potential differences in the biological functions of genes among high and low-risk groups, GSEA was performed based on the gene signatures using the R package ‘clusterprofiler’ (version 4.6.2).

### Construction and evaluation of CMB-Signature models for the prediction of molecular signature scores in TCGA-PRAD and independent hospital cohort

Random forest regression model (randomForest package in R, Version 4.6-14) and 5-fold cross-validation strategy were deployed to assess the predictive power of CMBs towards the molecular signatures on hold-out samples (i.e., the samples were not used during training) in TCGA-PRAD patients, and therefore constructed five models per signature. Spearman correlation was used to evaluate the performance of predicted scores compared to the ground truth (i.e., scores estimated by GSVA). During the CMB-Signature model application in an independent hospital cohort, the predicted score for a specific signature was defined as an average score from all five models corresponding to this signature.

### Cellular Morphometric Biomarker Risk Score (CMBRS) Construction and Patient Stratification in the TCGA-PRAD cohort

The construction of the cellular morphometric biomarker risk score (CMBRS) in the TCGA-PRAD cohort is defined below. The coefficients of the final CMBs as categorical variables were obtained from multivariate CoxPH regression analysis on PFS:

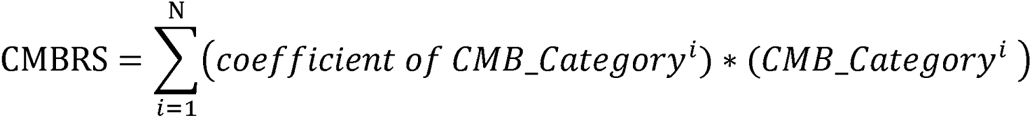

In the above equation, N is the number of final predictive CMBs that were pre-selected from NCT02430480 cohort, and *CMB_Category*^l^ is the category of the *i*^th^ CMB (i.e., CMB-high=1; CMB-low=0) where TCGA-PRAD cohort was divided into CMB-high/-low groups based on each CMB (cut-off estimated using survminer package in R, version 0.4.8, based on PFS). Then, we stratified the TCGA-PRAD cohort into three groups (High: top third; Intermediate: middle third; and Low: bottom third) based on CMBRS.

### Gene Risk Score (GERS) Construction and Patient Stratification in TCGA-PRAD cohort

The construction of the GERS in the TCGA-PRAD cohort is defined below. The coefficients of the genes as categorical variables were obtained from multivariate CoxPH regression analysis on PFS:

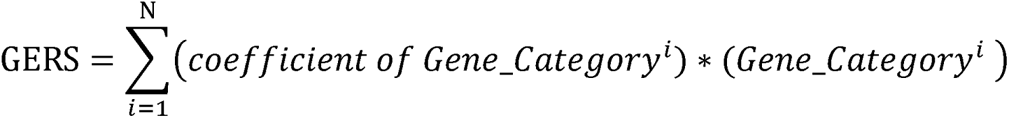

In the above equation, N is the number of genes that were pre-selected from NCT02430480 cohort that associated with specific SNPs, and *Gene_Category*^l^ is the category of the *i*^th^ gene (i.e., Gene-high=1; Gene-low=0) where TCGA-PRAD cohort was divided into Gene-high/-low groups based on each gene (cut-off estimated using survminer package in R, version 0.4.8, based on PFS). Then, we stratified the TCGA-PRAD cohort into three groups (High: top third; Intermediate: middle third; and Low: bottom third) based on GERS.

### Cellular Morphometric Biomarker and Gene Risk Score (CMBGERS) Construction and Patient Stratification in TCGA-PRAD cohort

Construction of the cellular morphometric biomarker and gene risk score (CMBGERS) in the TCGA-PRAD cohort is defined below. The coefficients of the CMBRS and GERS were obtained from multivariate CoxPH regression analysis on PFS:

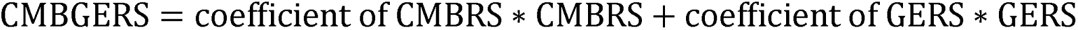

Then, we stratified the TCGA-PRAD cohort into three groups (High: top third; Intermediate: middle third; and Low: bottom third) based on CMBGERS.

### Immunohistochemical (IHC) Staining

IHC staining was carried out on 4-μm sections of formalin-fixed and paraffin-embedded tissues according to the standard protocol on a subset of the independent hospital cohort (20 patients in total; 10 patients from the ER-like group, and 10 patients from the INR-like group). The selection criteria are: (1) availability of sufficient specimens from needle biopsy before NADT for IHC staining; (2) availability of matching diagnostic slides from prostatectomy after NADT; and (3) balanced number of patients in ER-like and INR-like groups that meet the requirements (1) and (2). During IHC staining, sections were dewaxed and rehydrated in serial alcohol washes, and then endogenous peroxidase activities were blocked. After the nonspecific sites were saturated with 5% normal goat serum, the sections were incubated overnight at 4°C with anti-phospho-mTOR (1:50, mouse mAb, #67778-1-Ig, Proteintech) and anti-phospho-p70s6k (1:50, rabbit pAb, #ab2571, Abcam), and then incubated with anti-rabbit or anti-mouse Ig secondary antibodies. The sections were visualized using the biotin-peroxidase complex and counterstained with hematoxylin.

To assess p-mTOR and p-p70s6k, the stained sections were screened at low-power magnification (×20), and five hot spots were selected. The expression levels of p-mTOR and p-p70s6k were quantified using the H-score method (78), which was a semi-quantitative assessment combining both staining intensity (0: no staining; 1: weak staining; 2: moderate staining; and 3: intense staining) and percentage of positive cells with a numerical range from 0 to 300.

### Statistical analysis

All analysis was performed with R (Version 4.0.2). A predictive model based on CMB signature was constructed using logistic regression in R. Predictive power was assessed by accuracy, sensitivity, specificity, and area under the ROC Curve (AUC, pROC package in R, version 1.18.0). Survival differences between subtypes or groups were examined by log-rank test. Differences concerning the expression of immune checkpoints, immune cell infiltration, and genomic instability between groups were analyzed with the Mann-Whitney non-parametric test (for continuous variables) or Chi-square test (for categorical variables). P value (FDR corrected if applicable) < 0.05 was considered statistically significant.

## Data Availability

Whole slide images and clinical data of the NCT02430480 cohort were downloaded from the Cancer Imaging Archive (TCIA, https://doi.org/10.7937/TCIA.JHQD-FR46). Clinical and metadata were acquired from the original publication of the NCT02430480 study. Whole slide images of the TCGA-PRAD cohort were downloaded from the TCGA GDC portal (https://portal.gdc.cancer.gov/). Clinical and molecular data were downloaded from cBioportal (https://www.cbioportal.org/). All NCT02430480 and TCGA data were publicly available without modification. Raw data from the Nanjing Drum Tower Hospital is not currently permitted in public repositories because ethical and legal implications are still being discussed at an institutional level.

## Author Contributions

SG, KYJ, HC, and JHM conceptualized and designed the study. YH and YF led the independent hospital validation. AWM, MME, JHM, and HC performed formal analysis of public cohorts. YH and DL processed the raw data and performed formal analysis on the hospital validation cohort. SG, KYJ, KAI, DWW, RH, HC, JHM, MJPB, AJN, JPL, YF, HY, SP, WDC, and MB-H were involved in the critical review of the data and/or interpretation of results. AWM, MME, DWW, RH, HC, YH, DL, and JHM created the manuscript figures and supplementary materials. SG, KYJ, HC, YH, YF, KAI, and JHM drafted the manuscript. All authors edited, reviewed, revised, and approved the manuscript text. JHM and HC acquired funding for the study.

## Acknowledgment

This work was supported by the Department of Defense (DoD) BCRP, No. BC190820; and the National Cancer Institute (NCI) at the National Institutes of Health (NIH), No. R01CA184476. Lawrence Berkeley National Laboratory (LBNL) is a multi-program national laboratory operated by the University of California for the DOE under contract DE AC02-05CH11231. SG is supported by the NCI, award no. K08CA273542. Perez-Losada’s lab is sponsored by Grant PID2020-118527RB-I00 funded by MCIN/AEI/10.13039/501100011039; Grant PID2023-153081OB-I00 funded by MCIN/AEI/10.13039/501100011039” the Regional Government of Castile and León (CSI144P20).

